# Offset analgesia and onset hyperalgesia with different stimulus ranges

**DOI:** 10.1101/2020.06.01.20113613

**Authors:** Jens Fust, Maria Lalouni, Viktor Vadenmark Lundqvist, Emil Wärnberg, Karin B. Jensen

## Abstract

Offset analgesia (OA), a large reduction in pain following a brief increase in intensity of an otherwise stable painful stimulus, has been established by a large body of research. But the opposite effect, onset hyperalgesia (OH), a disproportional hyperalgesic response following a briefly decreased intensity of a painful stimulus, has only been investigated in one previous study. The aim of this study was to induce OA and OH in healthy participants and explore the effects of different stimulus ranges on OA and OH. A total of 62 participants were tested in two identical experiments. OA and OH conditions included two different temperature deviations (±1°C/±2°C) from baseline and were compared to a constant temperature (control). OA was successfully elicited in three out of four conditions. A dose-response relationship was demonstrated between the increase of temperature and the resulting hypoalgesic response. OH was only elicited in one out of four conditions (OH_2°C_ in experiment 1). Exploratory analysis showed that OA and OH responses were only weakly correlated. The asymmetry between pain responses following a brief temperature increase and decrease could be seen as evidence for different mechanisms involved in the pain responses to increasing and decreasing temperature. This asymmetry may also be explained by high temperatures in OA condition (+1°C/+2°C above baseline) that could be seen as salient “learning signals”, which augment the response to following changes in temperature.

## INTRODUCTION

The ability to feel pain is essential for our survival as it provides a warning signal for potential tissue damage. Yet, the perception of pain varies as a result of contextual adaptation. For example, pain intensity can be both upregulated and downregulated based on the temporal characteristics of a noxious stimulus. Evidence suggest that a noxious stimulus with fast onset is experienced as more painful than a stimulus with slow onset.^1^ In terms of the motivational aspects of pain, increasing painful stimulation may represent a strong signal of imminent tissue damage, and the opposite, decreasing painful stimulation, can be seen as a safety signal.^2^ Offset analgesia (OA) is an example of a manipulation of the temporal signature of noxious stimulation associated with hypoalgesia.^3^ More specifically, OA represents a disproportional reduction of pain following a brief increase and decrease of an otherwise stable painful stimulus.

The function of OA is not entirely clear, even if it has been described in terms of a temporal contrast enhancement mechanism that amplifies changes in the afferent signal.^4, 5^ OA is also associated with biological correlates related to central pain modulation, such as neural activation at the spinal level and the Periaqueductal Grey (PAG).^6, 7^ In contrast to other experimental paradigms that measure central pain modulation, such as temporal summation and conditioned pain modulation, the OA effect has not been suppressed by pharmacological blocking of specific neurotransmitter receptors.^8, 9^

OA demonstrates that fluctuating heat can produce *hypo*algesia, but less effort has been put on examining the opposite response, *hyper*algesia. Using a simple 2-step sequence of temperatures on the skin (48°C to 49°C and 49°C to 48°C), compared to constant heat, Mørch et al. demonstrated a disproportional decrease and increase of pain, respectively.^10^ In a recent study, Alter et al. produced “onset hyperalgesia” (OH) by inverting the standard OA 3-step sequence.^11^ Even though the evidence is sparse, these studies indicate the existence of a bi-directional temporal contrast mechanism, that can both amplify and weaken the pain response to dynamic noxious stimulation.

The aim of experiment 1 was to elicit hyperalgesia with an OH paradigm and compare to the pain response elicited by the standard OA design. To further study the effects of temperature changes on hyperalgesia and hypoalgesia, we also included two different temperature ranges for both the OA and OH sequence, resulting in four experimental conditions (OA_1°C_, OA_2°C_, OH_1°C_, OH_2°C_), as well as a control condition with constant temperature. We hypothesized that hyperalgesic response could be produced using the OH design, as well as hypoalgesic responses using the standard OA design. Moreover, we hypothesized that a larger temperature range would produce larger hyperalgesic and hypoalgesic responses. Experiment 2 was a direct replication of experiment 1. The procedure was identical to experiment 1, but more participants were included in order to increase statistical power. Also, both men and women were tested (all women in experiment 1). Because the method of statistical analysis was decided post-hoc in experiment 1, experiment 2 was conducted in order to corroborate the results of the first study.

## METHODS

### Participants

A total of 63 healthy participants were recruited for the study. One participant was excluded from experiment 2 because the participant did not look at the NRS scale during pain stimulation. Twenty-one women (mean age: 24 ± 2.7 years) were included in experiment 1 and 41 participants (22 women; mean age: 25 ± 4.5 years) in experiment 2. All participants were recruited through advertisement on university campuses in Stockholm and on the internet. The regional Ethics Review Board in Stockholm approved the study (Dnr: 2018/1367-31/1) and all subjects gave written informed consent.

### Procedure

Experiment 1 and 2 followed the same procedure. Heat stimuli were administered with a thermal stimulator (Somedic Senselab AB, Hörby, Sverige). Temperature increased and decreased at a rate of 5°C/s. A 30×30 mm thermal probe was attached to participants’ left calf. Participants used a trackball to continuously rate their pain intensity on a numeric rating scale (NRS) that was displayed on a screen, ranging from 0 (no pain) to 10 (worst imaginable pain). Individual pain sensitivity was calibrated before the experiment with 5 second heat stimuli ranging from 38 to 50°C. Participants were then given an additional 15 seconds heat stimulation set to each individual’s 5 NRS, predicted from the calibration data. If the pain rating ranged between 4 and 6 NRS this temperature was used as the baseline temperature in the experimental phase, otherwise the procedure was repeated with a higher or lower temperature until the desired pain rating was reached. OA and OH protocols were conducted with two different stimulus ranges during the mid-phase (T2) of the heat stimulation, with either a 1°C or 2°C increase/decrease (OA_1°C_, OA_2°C_, OH_1°C_, OH_2°C_). We decided to use a slightly modified version of the OH design used by Alter et al.^11^ In addition to using two different temperature ranges we decided to keep the individually calibrated baseline temperature constant in all conditions, only varying the temperature during the second phase of the procedure. For example, if the baseline temperature was set to 48°C, the participant would be exposed to the following temperature sequences: OA_1°C_, 48-49-48; OA_2°C_, 48-50-48; OH_1°C_, 48-47-48; OH_2°C_, 48-46-48; and control (constant temperature), 48-48-48. A constant temperature (no increase/decrease) was used as control. The presentation order of the conditions was randomized and each condition was followed by a 50 s break with a stand-by temperature of 38°C.

### Statistical analysis

The same statistical analyses were used for both experiments. First, we calculated mean pain ratings for each condition during the last 13 seconds of stimulation (from the time when the ratings started to diverge in T3 until temperature started to return to baseline). This time window was determined by examining the data in experiment 1. Second, we performed two repeated measures 1 × 3 ANOVA on these mean pain ratings; one including the offset conditions and the control condition (OArc, OA_2°C_, control) and one including the onset conditions and control condition (OH_1°C_, OH_2°C_, control). Third, if the ANOVA models reached statistical significance, we performed paired t-tests to compare the pain ratings between individual conditions. Corrections for multiple tests were done using the Benjamini-Hochberg method.^12^ A hypoalgesic response was determined as a difference between OA and the control condition, and a hyperalgesic response as a difference between OH and the control condition. Last, we conducted an exploratory analysis to examine the level of symmetry between OA and OH responses. Data from experiment 1 and 2 were merged. To be able to compare OA and OH, we subtracted the control conditions from each experimental condition, and inverted the OA conditions. Then we calculated subtracted offset effects and onset effects for each participant and stimulus range. The subtracted offset effect and onset effects was defined as the difference between the minimum rating during T2 (9 to 20 sec) and maximum rating during T3 (20-33 sec). Finally, we calculated Pearson correlation coefficients between the offset effect and the onset effect for each stimulus range. All calculations were made using Python 3.7.5. Repeated measures ANOVA was calculated using AnovaRM from the Python library statsmodels 0.10.1.^13^

## RESULTS

In experiment 1, we found a significant main effect of condition on pain ratings in the OA-model, *F*(2, 40) = 10.86, *p* < .001. Post-hoc tests showed that there was a significant hypoalgesic response during OA_2°C_, *t*(20) = 4.57, *p* < .001, but not during OArc. The hypoalgesic response was stronger in OA_2°_ compared to OA_1°C_, *t*(20) = 3.15, *p* = .008. We also found a significant main effect of condition on pain ratings in the OH-model, *F*(2, 40) = 3.72, *p* = .033. Post-hoc tests revealed a significant hyperalgesic response during OH_2°C_, *t*(20) = −3.19, *p* = .007, but not during OH_1°C_, and no significant difference between OH_2°C_ and OH_1°C_.

In experiment 2, we found a significant main effect of condition on pain ratings in the OA-model, *F*(2, 80) = 39.10, *p* < .001. Post-hoc tests showed that there was a significant hypoalgesic effect during both OA_1°C_, *t*(40) = 5.39, *p*<.001 and OA_2°C_, *t*(40) = 6.97, *p*<.001. As in the first experiment the hypoalgesic effect was stronger during OA_2°_ than OA_1°C_, *t*(40) = 4.51, *p* < .001. The main effect of condition on pain ratings in the OH-model was not significant, F(2, 80) = 3.72, *p* = .050.

In the exploratory analysis we found small but significant correlations between the offset and the onset effect with ±1°C stimulus range (*r*(60) = .27, *p* = .031) and between the offset and the onset effect with ±2°C stimulus range (*r*(60) = .27, *p* = .029).

## DISCUSSION

The aim of these two experiments was to examine if a hyperalgesic OH response could be induced by an inverted version of a well-documented hypoalgesic OA paradigm, and to determine if different stimulus ranges affect the OA and OH responses. Here, OA was successfully elicited in three out of four conditions. We also demonstrated that a larger stimulus range (i.e., larger increase/decrease in temperature), produces larger OA effects. The latter result indicates that there is a dose-response relationship between the increase of temperature and the following hypoalgesic response. Moreover, this result also indicates that a ±2°C design could be superior to a ±1°C design in studies where statistical power is an issue, for example studies of clinical populations with small samples size and/or between-group comparisons.

OH was only induced in one out of four conditions (OH2°c in experiment 1). Although these results are far from conclusive, it is possible that OH is a less stable phenomenon than OA. At first glance this might seem surprising as one may think that a responsive pain system would yield significant results in both the hyper- and hypoalgesic direction. Yet, we found that OA was more reliably induced than OH. The weak correlation between the OA and OH effects in the exploratory analysis further highlights the asymmetry between OA and OH. Even though Alter et al. emphasized the similarities between OA and OH in their study, they did report measurements of the OA and OH effects that were only weakly correlated.^11^ Mørch et al. also found that decreases in noxious temperature leads to slower but larger changes in pain compared to increases in temperature, which led them to propose that there are different mechanisms underlying pain responses to increases and decreases of temperature.^10^ The weak correlation between OA and OH effects found in our study support the notion of dual mechanisms.

A predictive coding perspective could also be useful in understanding the asymmetric response to rises and falls of noxious heat that we observed in the study. One important difference between the OH and OA conditions is that the latter involves temperatures 1°C or 2°C above baseline (calibrated as 5 out of 10 NRS). The brief but sharp rise in temperature in the OA conditions, can be seen as a salient “learning signal” that affects pain modulation upon return to baseline. As the noxious input during the short increase of temperature deviates from the predicted sensation, an error signal may feed forward to adjust the perception and/or update the relevant generative models^14^. Hence, the mismatch between top-down predictions of perceived pain, and bottom-up noxious signals, provides a mechanism for pain adaptation. In the case of OA this adaptation is expressed as inhibitory modulation of noxious heat. However, in the case of OH it is unclear if the brief decrease in temperature (−1°C or −2°C) may create a similarly salient surprise and motivate a subsequent adjustment of pain perception. This could also explain the discrepancy between our results and the previous study of OH. While our OH design used a baseline temperature (during T1 and T3) calibrated to each participants’ pain level of 5 NRS, Alter et al.^11^ used a 5 NRS + 1°C as baseline temperature, which could result in a “saliency-matched” learning signal for the OA and OH conditions, explaining the discrepancy between our and their finding.

In conclusion, the results from the present study provide evidence for dose-response pain regulation in response to the OA paradigm, and highlight the motivational role of the learning-signal in temporal contrast enhancement of pain. Future studies should determine it OA and OH represent dual mechanisms, or if temporal contrast enhancement is symmetric.

## Data Availability

Data will be made available upon request

## FIGURES

**Figure 1.**
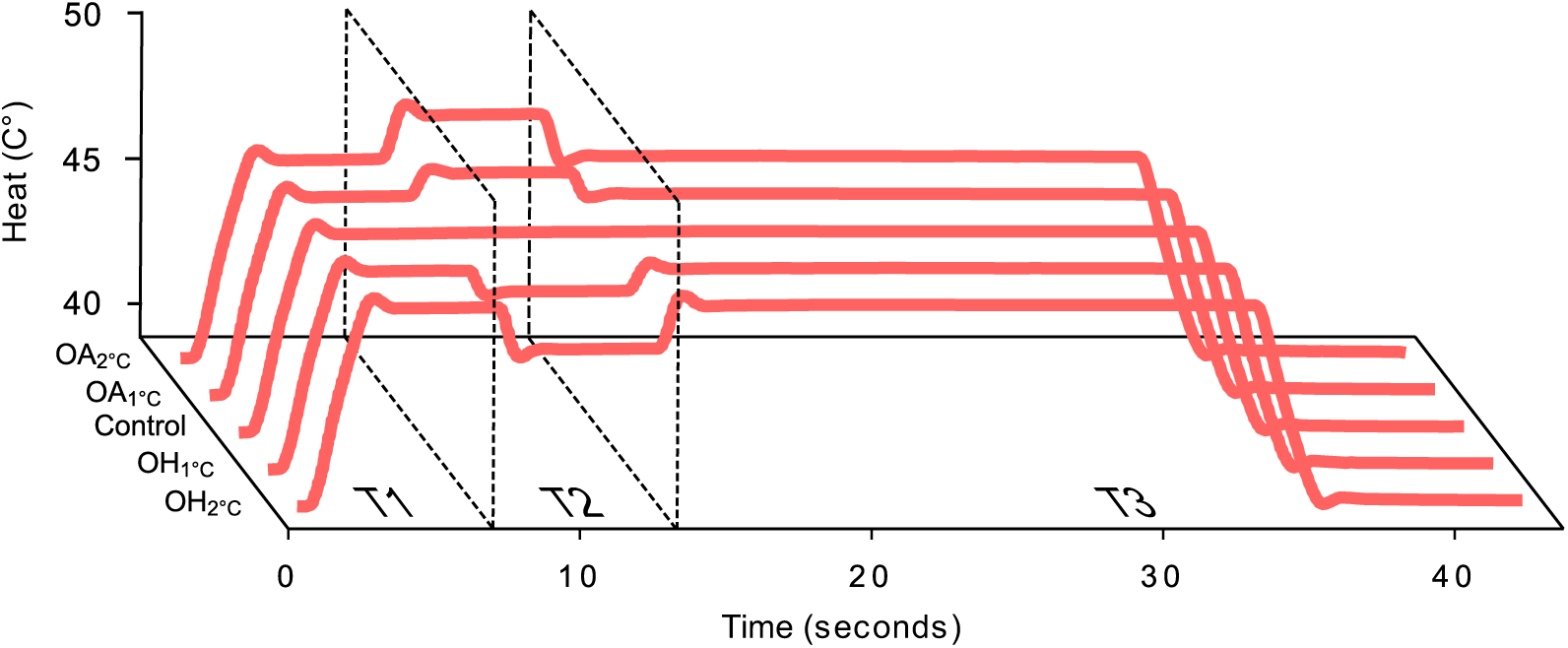
Mean heat stimulation (°C) during the four OA/OH conditions and the control condition. T1, T2 and T3 refers to three time intervals and denotes T1=baseline temperature pre manipulation, T2=temperature manipulation (±1°C or ±2°C), T3=baseline temperature post manipulation.

**Figure 2.**
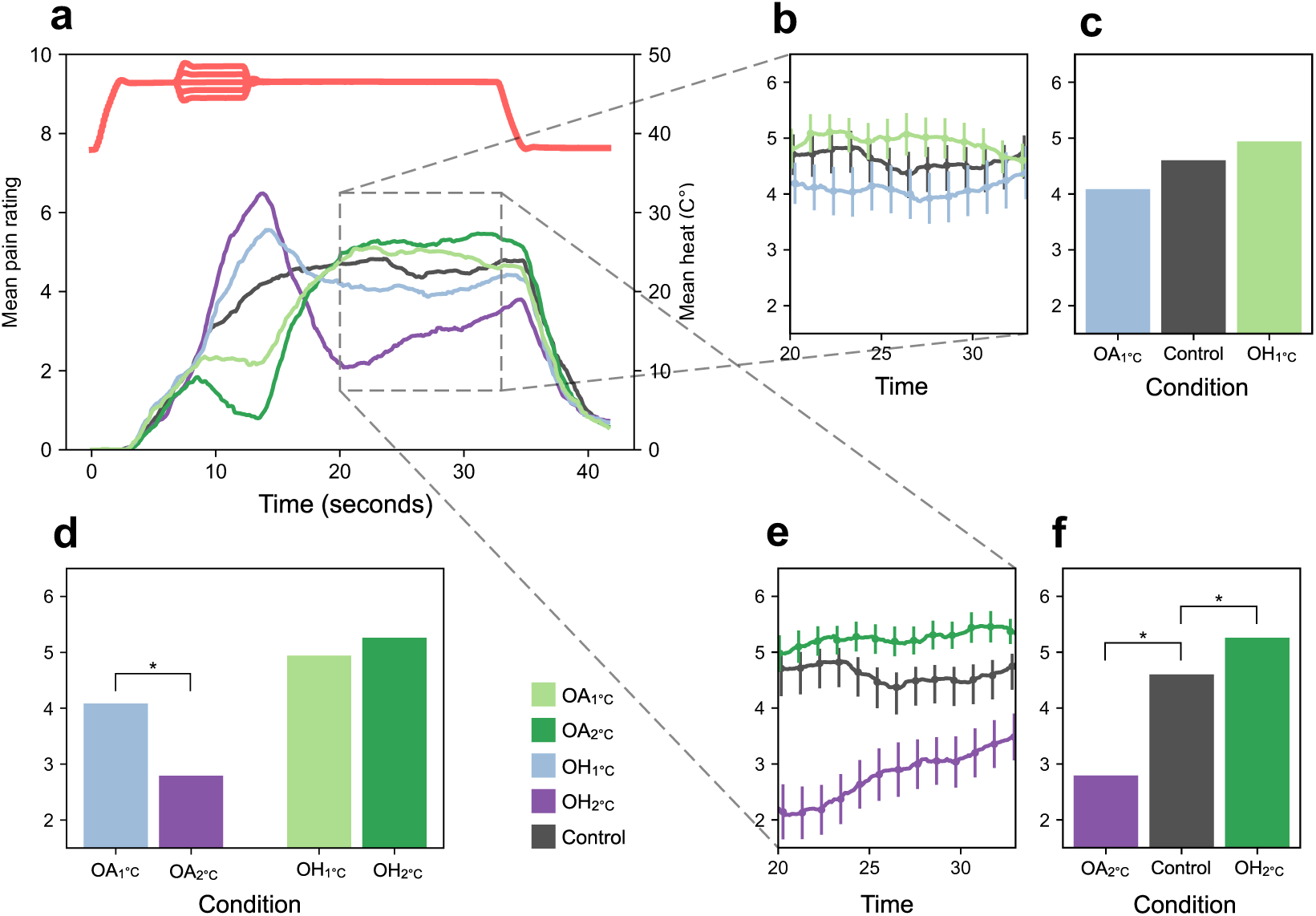
Results from experiment 1. **(a)** Mean pain ratings (left axis) and mean temperature (right axis) during OA 1°C, OA2C, OH1°C, OH2°c, and control condition **(b)** Mean pain ratings for ±1°C conditions and Control during the last 13 seconds of heat stimulation (error bars: ±1 within-subject SE). **(c)** Comparison between pain ratings for ±1°C conditions and Control during the last 13 seconds of heat stimulation. **(d)** Comparisons between pain ratings between ±1°C and +2°C conditions during last 13 seconds of heat stimulation. **(e)** Mean pain ratings for ±2 conditions and control during the last 13 seconds of heat stimulation (error bars: ±1 within-subject SE). **(f)** Total mean pain ratings for ±1 conditions and control condition during the last 13 seconds of heat stimulation

**Figure 3.**
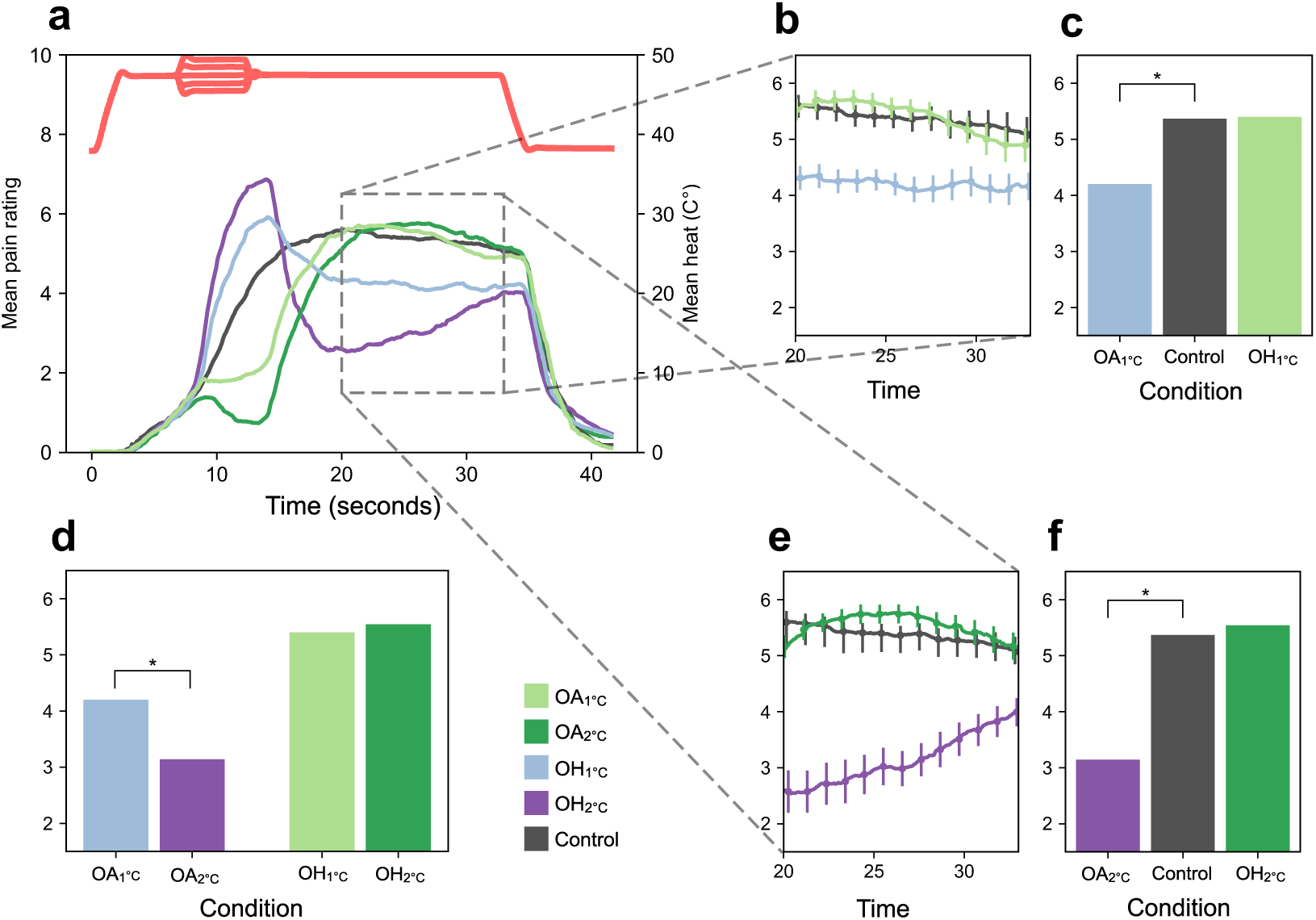
Results from experiment 2. **(a)** Mean pain ratings (left axis) and mean temperature (right axis) during OArc, OA2C, OH1°c, OH2°c, and control condition **(b)** Mean pain ratings for ±1°C conditions and control during the last 13 seconds of heat stimulation (error bars: ±1 within-subject SE). **(c)** Comparison between pain ratings for ±1°C conditions and control during the last 13 seconds of heat stimulation. **(d)** Comparisons between pain ratings between±1°C and +2°C conditions during last 13 seconds of heat stimulation. **(e)** Mean pain ratings for ±2 conditions and control during the last 13 seconds of heat stimulation (error bars: ±1 within-subject SE). **(f)** Total mean pain ratings for ±1 conditions and control condition during the last 13 seconds of heat stimulation

**Figure 4.**
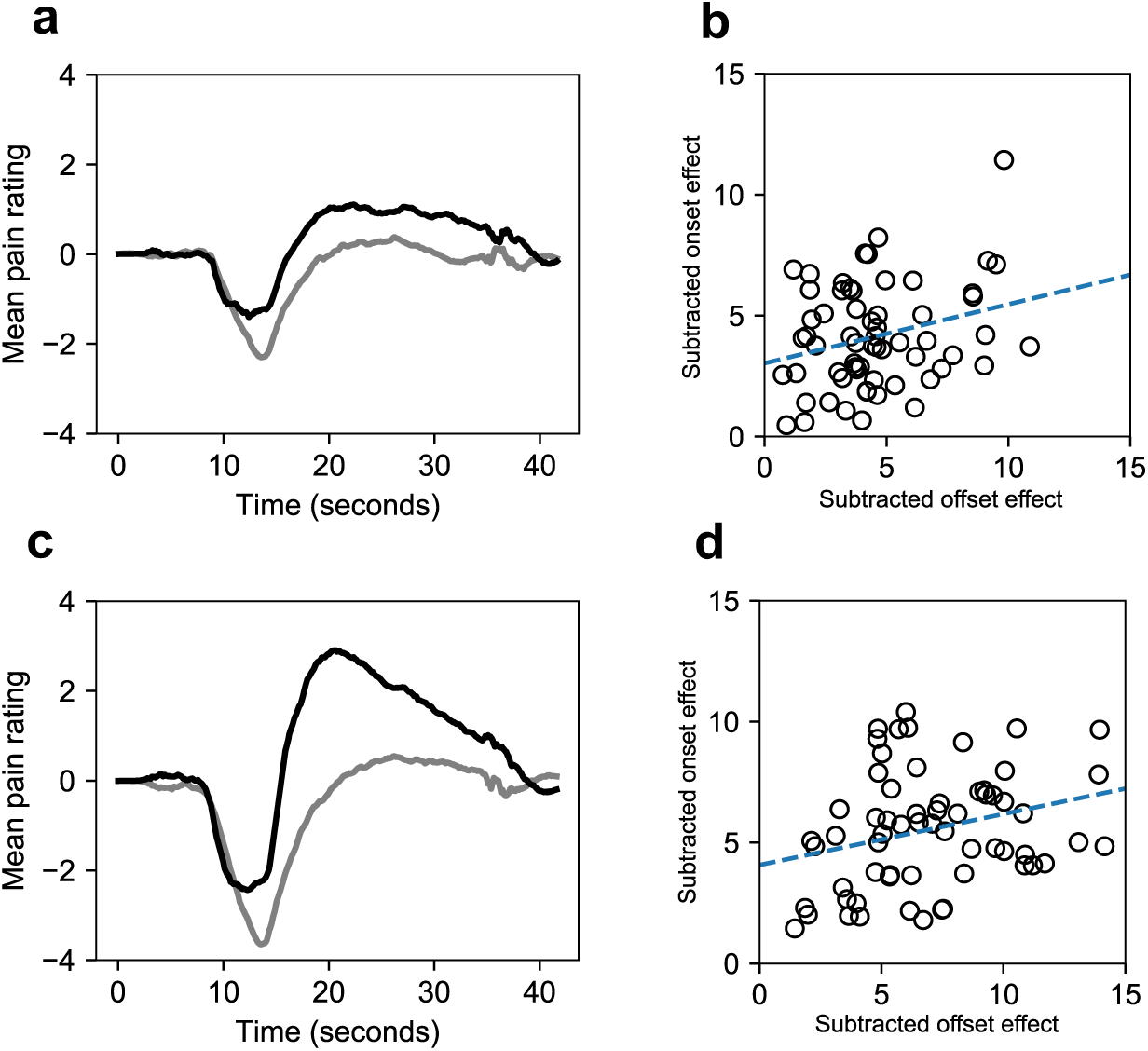
Results from exploratory analysis of OA OH symmetry. **(a)** The grey line represents the difference between pain ratings during OH1°c and control. The black line represents the inverted difference between pain ratings during OA1°c and control. **(b)** Correlation between subtracted offset effect and subtracted onset effect during ±1°C conditions. **(c)** The grey line represents the difference between pain ratings during OH2°c and control. The black line represents the inverted difference between pain ratings during OA2°c and control. **(d)** Correlation between subtracted offset effect and subtracted onset effect during ±2°C conditions.

## REFERENCES

1. Yarnitsky D, Ochoa JL. Studies of heat pain sensation in man: perception thresholds, rate of stimulus rise and reaction time: Pain. 1990;40(1):85–91. doi:10.1016/0304-3959(90)91055-N

2. Cecchi GA, Huang L, Hashmi JA, et al. Predictive dynamics of human pain perception. Morrison A, ed. PLoS Comput Biol. 2012;8(10):e1002719. doi:10.1371/journal.pcbi.1002719

3. Grill JD, Coghill RC. Transient analgesia evoked by noxious stimulus offset. J Neurophysiol. 2002;87(4):2205–2208. doi:10.1152/jn.00730.2001

4. Yelle MD, Rogers JM, Coghill RC. Offset analgesia: a temporal contrast mechanism for nociceptive information: Pain. 2008;134(1):174–186. doi:10.1016/j.pain.2007.04.014

5. Petre B, Tetreault P, Mathur VA, et al. A central mechanism enhances pain perception of noxious thermal stimulus changes. Sci Rep. 2017;7(1):3894. doi:10.1038/s41598-017-04009-9

6. Sprenger C, Stenmans P, Tinnermann A, Büchel C. Evidence for a spinal involvement in temporal pain contrast enhancement. NeuroImage. 2018;183:788–799. doi:10.1016/j.neuroimage.2018.09.003

7. Yelle MD, Oshiro Y, Kraft RA, Coghill RC. Temporal filtering of nociceptive information by dynamic activation of endogenous pain modulatory systems. J Neurosci. 2009;29(33): 10264–10271. doi:10.1523/JNEUROSCI.4648-08.2009

8. Martucci KT, Eisenach JC, Tong C, Coghill RC. Opioid-independent mechanisms supporting offset analgesia and temporal sharpening of nociceptive information: Pain. 2012;153(6):1232–1243. doi:10.1016/j.pain.2012.02.035

9. Niesters M, Proto PL, Aarts L, Sarton EY, Drewes AM, Dahan A. Tapentadol potentiates descending pain inhibition in chronic pain patients with diabetic polyneuropathy. Br J Anaesth. 2014; 113(1):148–156. doi:10.1093/bja/aeu056

10. Mørch CD, Frahm KS, Coghill RC, Arendt-Nielsen L, Andersen OK. Distinct temporal filtering mechanisms are engaged during dynamic increases and decreases of noxious stimulus intensity: Pain. 2015;156(10): 1906–1912. doi:10.1097/j.pain.0000000000000250

11. Alter BJ, Aung MS, Strigo IA, Fields HL. The onset and offset of noxious stimuli robustly modulate perceived pain intensity. *bioRxiv*. 2020. doi:10.1101/2020.03.18.996769

12. Benjamini Y, Hochberg Y. Controlling the false discovery rate: a practical and powerful approach to multiple testing. JR Stat Soc Ser B Methodol. 1995;57(1):289–300.

13. Seabold S, Perktold J. Statsmodels: Econometric and statistical modeling with python. In: Proceedings of the 9th Python in Science Conference. Vol 57. Scipy; 2010: 61.

14. Büchel C, Geuter S, Sprenger C, Eippert F. Placebo analgesia: a predictive coding perspective. Neuron. 2014;81(6):1223–1239. doi:10.1016/j.neuron.2014.02.042

